# Clinical effectiveness of drugs in hospitalized patients with COVID-19: a systematic review and meta-analysis

**DOI:** 10.1101/2020.09.11.20193011

**Authors:** R.A. Abeldaño Zuñiga, S.M. Coca, G.F. Abeldaño, R.A.M. González Villoria

**Affiliations:** Postgraduate Department. University of Sierra Sur. Mexico.; Public Health Research Institute. University of Sierra Sur. Mexico.; School of Medicine. University of Sierra Sur. Mexico.

**Keywords:** Drugs, Antivirals, Clinical improvement, Mortality, COVID-19, SARS-CoV2.

## Abstract

**Objective:** The aim was to assess the clinical effectiveness of drugs used in hospitalized patients with COVID-19 infection.

**Method:** We conducted a systematic review of randomized clinical trials assessing treatment with remdesivir, chloroquine, hydroxychloroquine, lopinavir, ritonavir, dexamethasone, and convalescent plasma, for hospitalized patients with a diagnosis of SARS-CoV-2 infection. The outcomes were mortality, clinical improvement, duration of ventilation, duration of oxygen support, duration of hospitalization), virological clearance, and severe adverse events.

**Results:** A total of 48 studies were retrieved from the databases. Ten articles were finally included in the data extraction and qualitative synthesis of results. The meta-analysis suggests a benefit of dexamethasone versus standard care in the reduction of risk of mortality at day 28; and the clinical improvement at days 14 and 28 in patients treated with remdesivir.

**Conclusions:** Dexamethasone would have a better result in hospitalized patients, especially in low-resources settings.

**Significance of results:** The analysis of the main treatments proposed for hospitalized patients is of vital importance to reduce mortality in low-income countries; since the COVID-19 pandemic had an economic impact worldwide with the loss of jobs and economic decline in countries with scarce resources.

## Introduction

In December 2019, the first cases of an emerging disease, currently called COVID-19, were presented. The spread of SARS-CoV2 was declared a pandemic in March 2019, which generated a global health emergency [1]. From the first cases, treatments based on drug repositioning were implemented [2].

The disease has different degrees of severity, having asymptomatic infected people, people with a mild disease without pneumonia, or mild pneumonia. The severe degree, with dyspnea, bradypnea, hypoxia, pulmonary infiltrates, and the critical clinical condition, with respiratory failure, septic shock, or multi-organ failure, requires optimal treatment and hospital care [3].

The fatality rates of this infection vary throughout the world, being higher in Africa, India, the USA, Mexico, and Brazil, where various comorbidities in the population such as hypertension, obesity, and diabetes increase fatality [4,5]. Despite implementing recommended control measures in Latin America, the countries have been affected differently, with high fatality rates related to the differences in health services in different countries [6].

The use of antivirals or other repositioning drugs is essential for clinical improvement and survival. In the absence of a specific treatment, in vitro and in vivo studies have been proposed to use existing drugs such as tocilizumab (monoclonal antibodies) [7], remdesivir (antiviral) [8], chloroquine and hydroxychloroquine (antimalarial) [9,10], lopinavir and ritonavir (antiretrovirals) [11], dexamethasone (glucocorticoid) [12], convalescent plasma (neutralizing antibody) [13], and traditional medicine [14,15]. All of them showed beneficial effects in preclinical studies and some clinical studies; however, evaluating the treatments used in hospitalized patients is required. The aim was to assess the clinical effectiveness of antivirals used in hospitalized patients with COVID-19 infection.

## Methods

A systematic review was carried out adhering to the PRISMA guidelines for conducting systematic reviews [16]. The question in this review was:

What is the clinical effectiveness of antivirals in hospitalized patients with COVID-19 infection? To conduct the review, the PICOS structure was followed according to the following points:

- Patients: Adults hospitalized with a diagnosis of SARS-CoV-2 infection.
- Intervention: Treatment with the following drugs: remdesivir, chloroquine, hydroxychloroquine, lopinavir, ritonavir, dexamethasone, and convalescent plasma.
- Comparison: Standard care or placebo.
- Outcomes: Early mortality, late mortality, 28 days mortality, clinical improvement at seven days, clinical improvement at 14 days, clinical improvement at 28 days, duration of ventilation (days), duration of oxygen support (days), duration of hospitalization (days), virological clearance, and severe adverse events.
- Studies (type of): Clinical trials published in peer-reviewed journals. The search was carried out in PubMed, Scopus, and Web of Science databases, between August 20th and September 9th, 2020. The references of the selected articles were also reviewed for an integral reading to include additional studies not indexed in these databases. The clinicaltrials.gov website was also scanned to obtain potential published reports of registered trials. The search strategies included the following keywords: remdesivir, chloroquine, hydroxychloroquine, lopinavir, ritonavir, dexamethasone, convalescent plasma, COVID-19, SARS-CoV2, and hospitalized. See the supplemental file for more details on the search strategies.

Studies that met the following criteria were included: I) Controlled clinical trials, II) Studies that included hospitalized patients with SARS-CoV-2 infection, III) Published in 2020, IV) Published in English, Chinese, Spanish or Portuguese. The exclusion criteria were: I) Not being a clinical trial, II) Not treating hospitalized patients.

All references were managed with Mendeley® software. The selection of the articles began with the removal of duplicate articles, and proceeded with the reading of the title and abstract, carried out independently by reviewers 1, 2, and 3. The final decision in cases of disagreement was based on the criteria of a fourth reviewer. In the second phase, the same reviewers read the full text of the studies to define which would be included for the extraction and synthesis of data. The data were stored in Microsoft Office Excel spreadsheets and organized in an instrument constructed by the authors considering: Characteristics of the study (author, year, country), sample, study design, and characteristics of the results. The risk of bias of the studies was evaluated using the ROB2 tool [17]. The included studies were independently assessed by reviewers 1 and 2 (See supplemental file).

The qualitative synthesis was developed following the assessed outcomes: Early mortality, late mortality, 28 days mortality, clinical improvement at seven days, clinical improvement at 14 days, clinical improvement at 28 days, duration of ventilation (days), duration of oxygen support (days), duration of hospitalization (days), virological clearance, and severe adverse events.

Finally, the meta-analysis of random effects was conducted for articles included assessing virological clearance at day 7, and meta-analysis of fixed effects was conducted for included articles assessing severe adverse events. The review protocol was registered on the PROSPERO platform (CRD42020184436).

## Results

A total of 48 studies were retrieved from the databases. After the removal of 6 duplicates, 42 articles were read in title and abstract. Twenty-seven were eliminated, resulting in 15 articles for full-text reading. Ten articles were finally included in the data extraction and qualitative synthesis of results (Figure 1).

**Figure 1.**
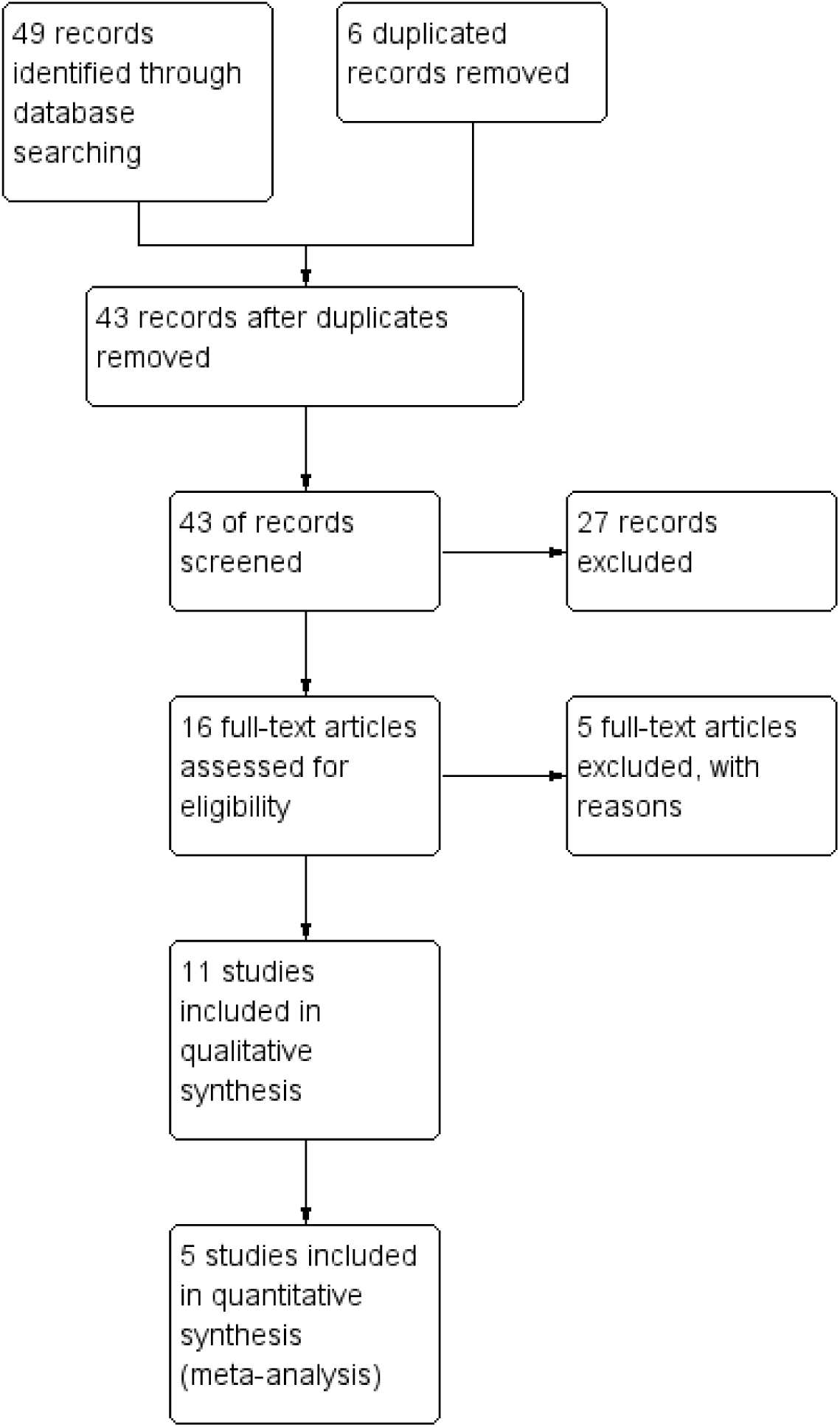
PRISMA flowchart of the inclusion process in the systematic review.

The overall risk of bias in the reviewed articles was established at low-risk in two studies [10,18]. The remaining eight studies were established at high risk or some concerns. More details can be seen in the supplemental file.

Two articles reported using lopinavir-ritonavir mixtures, two studies reported Remdesivir, three articles reported hydroxychloroquine, one study treated patients with chloroquine, two studies reported dexamethasone, and one study reported convalescent plasma. Patient samples ranged from 30 (the study with the fewest patients) to 6,425 (the study with the most patients). The retrieved results were: Early mortality (defined as mortality before 12 days), late mortality (defined as mortality after the 12th day), 28 days mortality, clinical improvement at 7, 14, and 28 days (defined by clinical scales), the mean duration of ventilation (in days), the mean duration of oxygen support (in days), the mean duration of hospitalization (in days), virological clearance (by laboratory tests), and severe adverse events (Table 1).

**Table 1.** Main characteristics of the included studies.

| Author | Study site | Design | Sample | Intervention | Control | Outcomes reported |
| --- | --- | --- | --- | --- | --- | --- |
| Cao 2020 | Hubei | Randomized, open-label, clinical trial | 99 intervention, 100 control | Lopinavir–Ritonavir | Standard care | Mortality at day 28, early mortality, late mortality, clinical improvement at day 7, 14, and 28, duration of ventilation, duration of oxygen support, duration of hospitalization, virological clearance, and adverse events |
| Hung 2020 | Hong Kong | Randomized, open-label, clinical trial | 86 intervention, 41 control | Lopinavir-Ritonavir-Ribavirin-Interferon Beta-1b | Lopinavir-Ritonavir | Mortality at day 28, clinical improvement at day 7, duration of hospitalization, virological clearance, and adverse events |
| Wang 2020 | Hubei | Randomized, double-blinded, clinical trial | 158 intervention, 79 control | Remdesivir | Placebo | Mortality at day 28, early mortality, late mortality, clinical improvement at day 7, 14, and 28, duration of ventilation, duration of oxygen support, duration of hospitalization, virological clearance, and adverse events |
| Chen 2020 | Shanghai | Randomized, open-label, clinical trial | 15 intervention, 15 control | Hydroxychloroquine | Standard care | Virological clearance and adverse events |
| Gautret 2020 | France | Non-randomized, open-label, clinical trial | 20 intervention, 16 control | Hydroxychloroquine | Standard care | Virological clearance |
| Tang 2020 | China | Randomized, open-label, clinical trial | 75 intervention, 75 control | Hydroxychloroquine | Standard care | Virological clearance and adverse events |
| Borba 2020 | Brazil | Randomized, double-blinded, clinical trial | 41 intervention, 40 control | Chloroquine High dosage | Chloroquine low dosage | Mortality at day 28, early mortality, and adverse events |
| RECOVERY 2020 | United Kingdom | Randomized, open-label, clinical trial | 2104 intervention, 4321 control | Dexamethasone | Standard care | Mortality at day 28 |
| Li 2020 | Hubei | Randomized, open-label, clinical trial | 52 intervention, 50 control | Convalescent Plasma | Standard care | Mortality at day 28, clinical improvement at day 7, 14, and 28, virological clearance, and adverse events |
| Spinner 2020 | US | Randomized, open-label, clinical trial | 197 intervention A, 199 intervention B, 200 control | Remdesivir | Standard care | Clinical improvement at day 7, 14, and 28, and adverse events |
| Tomazini 2020 | Brazil | Randomized, open-label, clinical trial | 151 intervention 148 control | Dexamethasone | Standard care | Mortality at day 28, and adverse events |

### 28 days mortality

Six clinical trials assessed the mortality of hospitalized patients at day 28 [10,18– 22], and one study reported mortality at day 30 [23]. The drugs applied as an intention of treatment reporting mortality were: lopinavir-ritonavir [19], lopinavir-ritonavir-ribavirin-interferon Beta-1b [23], remdesivir [18], chloroquine at high doses (600 mg) [10], dexamethasone [20,22], and convalescent plasma [21] (Table 2).

**Table 2.** Reported outcomes in the included studies.

| Author | Intervention (I) | Mortality at day 28<br>%(I)-%(C) | Early mortality<br>%(I)-%(C) | Late mortality<br>%(I)-%(C) | Clinical improvement at day 7<br>%(I)-%(C) | Clinical improvement at day 14<br>%(I)-%(C) | Clinical improvement at day 28<br>%(I)-%(C) | Duration of ventilation<br>Median (I)-(C) | Duration of oxygen support<br>Median (I)-(C) | Duration of hospital stay days<br>Median (I)-(C) | Virological clearance<br>%(I)-%(C) | adverse events<br>%(I)-%(C) |
| --- | --- | --- | --- | --- | --- | --- | --- | --- | --- | --- | --- | --- |
| Cao 2020 | Lopinavir – Ritonavir | 19.2% - 25% | 19% - 27.1% | 19.3% - 23.1% | 6.1% - 2% | 45.5% - 30% | 78.8% - 70% | 4 - 5 | 12 - 13 | 14 - 16 | 60.3% - 58.6% (day 28) | 20% - 32.3% |
| Hung 2020 | Lopinavir - Ritonavir - Ribavirin - Interferon Beta-1b | 0% - 0% | - | - | 4 - 8 (days to NEWS2 = 0) | - | - | - | - | 9 - 14.5 | 8 - 13 (median days) | 0% - 2% |
| Wang 2020 | Remdesivir | 14% - 13% | 11% - 15% | 14% - 10% | 3% - 2% | 27% - 23% | 65% - 58% | 7 - 15.5 | 19 - 21 | 25 - 24 | 75.6% - 83.1% (day 28) | 18% - 26% |
| Chen 2020 | Hydroxychloroquine | - | - | - | - | - | - | - | - | - | 86.7% - 93.3% (day 7) | 26.7% - 20% |
| Gautret 2020 | Hydroxychloroquine | - | - | - | - | - | - | - | - | - | 70% - 12.5% (day 6) | - |
| Tang 2020 | Hydroxychloroquine | - | - | - | - | - | - | - | - | - | 70.7% - 74.7% (day 28) | 3% - 0% |
| Borba 2020 | Chloroquine High dosage | - | 39% /- 15% | - | - | - | - | - | - | - | - | 18.9% - 11.1% |
| RECOVERY 2020 | Dexamethasone | 22.9% - 25.7% | - | - | - | - | - | - | - | - | - | - |
| Ling 2020 | Convalescent Plasma | 15.7% - 24% | - | - | 9.6% - 9.8% | 32.7% - 17.6% | 51.9% - 43.1% | - | - | - | 87.2% - 37.5% (day 3) | 3.8% - 0% |
| Spinner 2020 | Remdesivir | - | - | - | 48%*<br>56%**<br>47%*** | 77%*<br>76%**<br>68%*** | 90%*<br>90%**<br>83%*** | - | - | - | - | 5%*<br>5%**<br>9%*** |
| Tomazini 2020 | Dexamethasone | 56.3% - 61.5% | - | - | - | - | - | - | - | - | - | 3.3% - 6.1% |

%(I): % in the Intervention group %(C) % in the control group \* Intervention A (10 days remdesivir), \*\*Intervention B (5 days remdesivir), \*\*\*Control

### Early mortality

The early mortality, measured as the death produced before 12 days from patients allocation, was reported by a study using lopinavir-ritonavir [19], one trial using remdesivir [18], and one trial using chloroquine at high doses (600 mg) [10] (Table 2).

### Late mortality

The late mortality, measured as the death produced after 12 days from patients allocation, was only reported by two studies, one of them using lopinavir-ritonavir [19], and the other one using remdesivir [18] (Table 2).

### Clinical improvement

The clinical improvement was measured using the National Early Warning Score (NEWS) 2 [24]. It is an aggregate scoring system including six physiological parameters: respiration rate, oxygen saturation, systolic blood pressure, pulse rate, level of consciousness, and temperature. Clinical improvement at day seven was reported by three studies [18,19,21], while three studies reported clinical improvement at days 14 and 28 [18,19,21]. One study reported the median time (in days) to reach a NEWS2 score of zero [23] (Table 2). The study published by Spinner [8] also reported clinical improvement at days 7, 14, and 28, but it is not declared which scale was used to assess the clinical improvement.

### Duration of ventilation

This outcome was measured as the median number of days of duration of mechanical ventilation. It was reported by two studies using lopinavir-ritonavir [19] and Remdesivir [18] (Table 2).

### Duration of oxygen support

Two studies measured this outcome as the need for oxygen support through the nasal duct or mask, high-flow oxygen, or non-invasive ventilation [18,19]. The duration of oxygen support was reported in median days (Table 2).

### Duration of hospital stay

This outcome was reported in median days by three studies using lopinavir-ritonavir [19], lopinavir-ritonavir-ribavirin-interferon Beta-1b [23], and remdesivir [18] (Table 2).

### Virological clearance

This outcome was measured as the respiratory tract sample that was positive on RT-PCR, and it was reported as the virus clearance in respiratory samples in days after the allocation. One study reported this outcome at day 3 [21], one study at day 6 [25], two studies at day 7 [9,26], two at day 28 [18,19], and one study reported as the median days to reach a zero viral load [23] (Table 2).

### Adverse events

In this review, the data were extracted from nine studies reporting any severe adverse events [8–10,18,19,21–23,26] (It must be noted that a patient can develop one or more than one adverse event). Severe (or serious) adverse events were extracted as dichotomous data (Table 2). Of the nine studies that reported adverse events, only one has recorded no adverse events in any patient undergoing the intervention with lopinavir-ritonavir-ribavirin-interferon Beta-1b [23]. It is necessary to highlight the incidence of adverse events in studies with lopinavir-ritonavir [19], hydroxychloroquine [9], remdesivir [18], and chloroquine [10].

The conclusions reported by seven studies suggest that there is no benefit with the use of lopinavir-ritonavir [19], remdesivir [8,18], hydroxychloroquine [9,26] and chloroquine at high dosages [10]. However, two studies reported that dexamethasone resulted in lower mortality at day 28 among patients with severe clinical conditions [20] and a higher mean number of days alive and free form mechanical ventilation [22]; both studies together make up a total sample of 6724 patients. Another trial suggests that triple viral treatment (lopinavir-ritonavir-ribavirin-interferon Beta-1b) was superior to lopinavir-ritonavir alone in a sample of 127 patients [23]. Finally, one study suggests that hydroxychloroquine is significantly associated with viral load reduction in a sample of 36 patients [25] (Table 3).

**Table 3.**
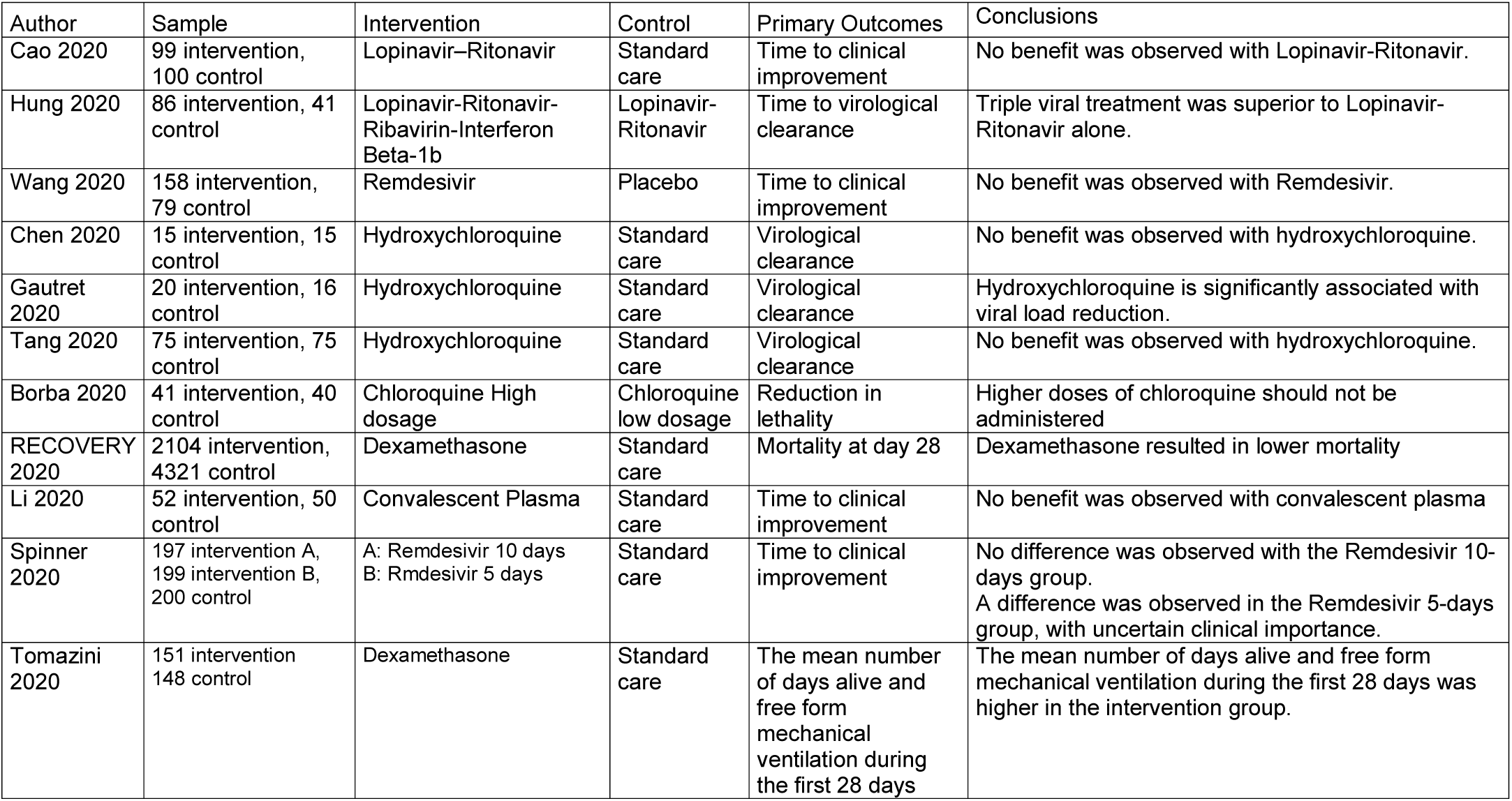
Primary outcomes and main conclusions of the included studies.

| Author | Sample | Intervention | Control | Primary Outcomes | Conclusions |
| --- | --- | --- | --- | --- | --- |
| Cao 2020 | 99 intervention, 100 control | Lopinavir–Ritonavir | Standard care | Time to clinical improvement | No benefit was observed with Lopinavir-Ritonavir. |
| Hung 2020 | 86 intervention, 41 control | Lopinavir-Ritonavir-Ribavirin-Interferon Beta-1b | Lopinavir-Ritonavir | Time to virological clearance | Triple viral treatment was superior to Lopinavir-Ritonavir alone. |
| Wang 2020 | 158 intervention, 79 control | Remdesivir | Placebo | Time to clinical improvement | No benefit was observed with Remdesivir. |
| Chen 2020 | 15 intervention, 15 control | Hydroxychloroquine | Standard care | Virological clearance | No benefit was observed with hydroxychloroquine. |
| Gautret 2020 | 20 intervention, 16 control | Hydroxychloroquine | Standard care | Virological clearance | Hydroxychloroquine is significantly associated with viral load reduction. |
| Tang 2020 | 75 intervention, 75 control | Hydroxychloroquine | Standard care | Virological clearance | No benefit was observed with hydroxychloroquine. |
| Borba 2020 | 41 intervention, 40 control | Chloroquine High dosage | Chloroquine low dosage | Reduction in lethality | Higher doses of chloroquine should not be administered |
| RECOVERY 2020 | 2104 intervention, 4321 control | Dexamethasone | Standard care | Mortality at day 28 | Dexamethasone resulted in lower mortality |
| Li 2020 | 52 intervention, 50 control | Convalescent Plasma | Standard care | Time to clinical improvement | No benefit was observed with convalescent plasma |
| Spinner 2020 | 197 intervention A, 199 intervention B, 200 control | A: Remdesivir 10 days<br>B: Rmdesivir 5 days | Standard care | Time to clinical improvement | No difference was observed with the Remdesivir 10-days group.<br>A difference was observed in the Remdesivir 5-days group, with uncertain clinical importance. |
| Tomazini 2020 | 151 intervention 148 control | Dexamethasone | Standard care | The mean number of days alive and free from mechanical ventilation during the first 28 days | The mean number of days alive and free from mechanical ventilation during the first 28 days was higher in the intervention group. |

## Meta-analysis

After discarding the individual articles that did not show conclusions in favor of the drugs used, five articles were included in the quantitative synthesis.

The result of two studies was integrated into the meta-analysis for comparing Dexamethasone versus standard care in the reduction of risk of mortality at day 28 [20,22]. This drug shows a benefit for patients in severe clinical conditions (OR: 0.86; IC: 0.76-0.96) (Figure 2).

**Figure 2.**
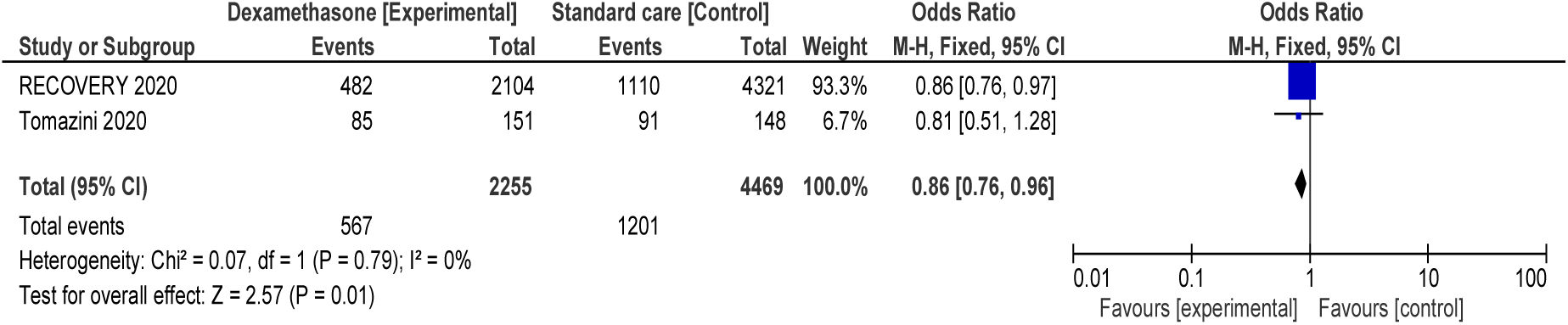
Forest plot of drugs for hospitalized patients with SARS-CoV2 infection. Comparison: Dexamethasone versus Standard care. Outcome: Mortality at day 28.

Two studies reporting remdesivir outcomes were compared to test the overall effect of this antiviral on clinical improvement on days 7, 14, and 28. The results show no association with clinical improvement at day 7 (OR: 1.03; IC: 0.70 - 1.51), but a very slight association with clinical improvement at day 14 (OR: 1.45; IC: 1.01- 2.08) and at day 28 (OR: 1.59; IC: 1.05- 2.38) (Figure 3). The drug was not associated with the presence of severe adverse events in the 10-days treatment group (OR: 0.57; IC: 0.36-0.92) (Figure S.1 in the supplemental file).

**Figure 3.**
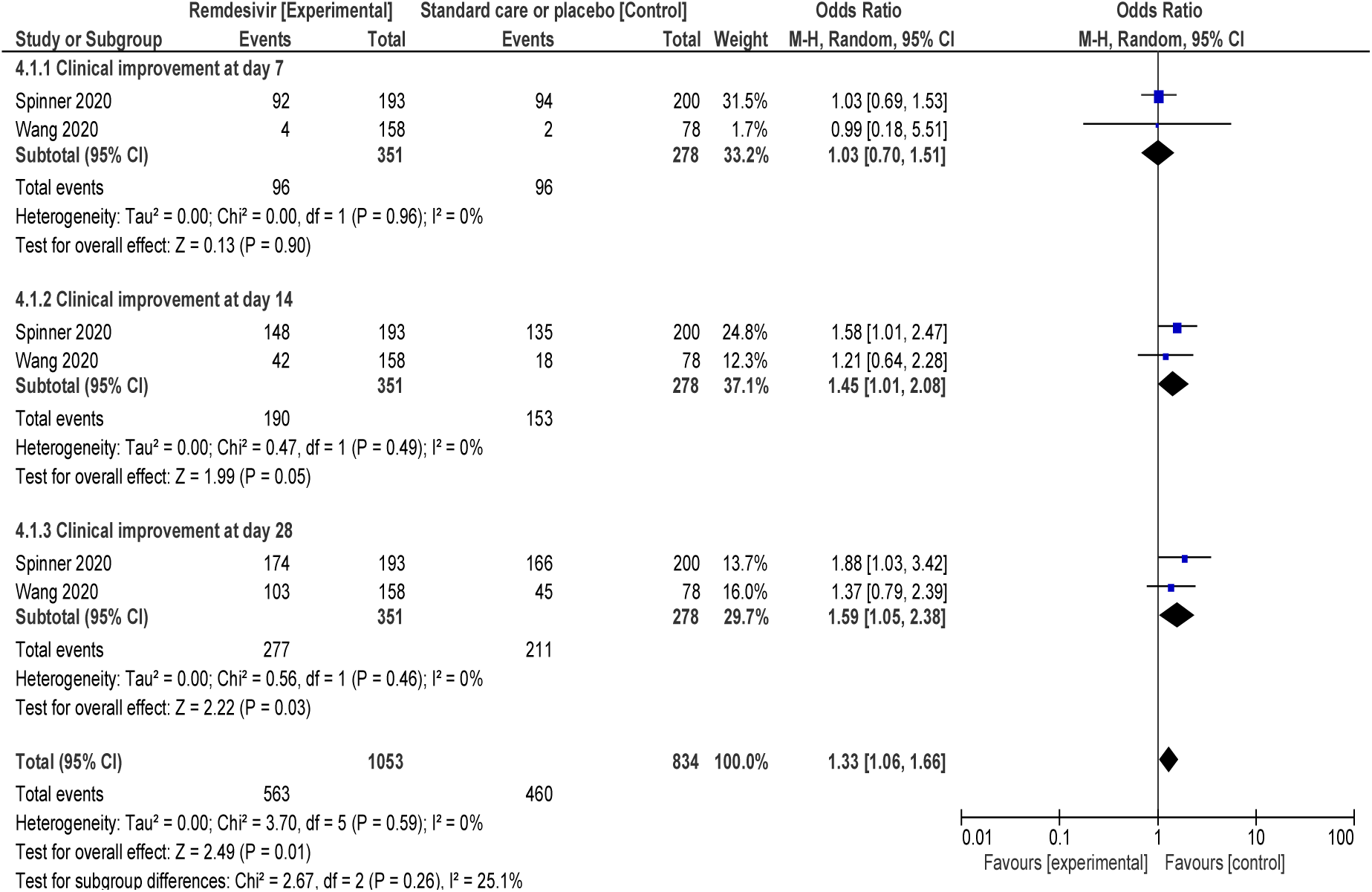
Forest plot of drugs for hospitalized patients with SARS-CoV2 infection. Comparison: Remdesivir versus Standard care or Placebo. Outcome: Clinical improvement

The results of 3 studies [9,25,26] were meta-analyzed to establish comparisons between the use of Hydroxychloroquine and Standard care, using the outcome "virological clearance at day 7". High heterogeneity was observed in the studies, so the meta-analysis of random effects suggests no benefits using this drug (Figure S.2 in the supplemental file). Also, the results of 2 trials [9,26] were meta-analyzed for the outcome of "severe adverse events". No heterogeneity was observed; therefore, a fixed-effects meta-analysis was run. The results show no differences in the risk of using the drug or the standard care (Figure S.3 in the supplemental file).

## Discussion

With the focus on adult hospitalized patients, this systematic review was able to identify nine clinical trials that were very heterogeneous among themselves, due to experimentation with different drugs and different administration regimens. In total, 8282 patients were included in hospitals in China, France, Brazil, the United Kingdom, and the United States.

Regarding the risk of bias of the included studies, it is essential to note that there were included eight with a high risk of bias or some concerns. The lack of blinding affected the risk of bias, mainly in studies launched under emergency conditions due to the international health crisis.

This study differs from another recent systematic review that evaluated antiviral drugs in patients with suspected, probable, or confirmed diagnosis of SARS-CoV2 infection [27]. Our study focuses only on hospitalized patients since, in some low-income Latin American countries, the epidemic has not yet reached its peak, and hospitals are experiencing saturation in their facilities [28–30].

The only drugs reported by more than one article published in peer-reviewed journals were hydroxychloroquine, remdesivir, and dexamethasone.

Hydroxychloroquine did not show benefits in virological clearance in our meta-analysis. Also, due to serious adverse events reported in another systematic review [27] have led to the conclusion that its use is not recommended. Regarding remdesivir, our meta-analysis has shown some association with clinical improvement on days 14 and 28. Furthermore, we observed a beneficial effect on adverse events in studies using remdesivir. Concerning mortality at day 28, dexamethasone has shown benefits in our meta-analysis in patients with severe clinical conditions.

In general, individual studies have concluded that no benefit was observed with lopinavir-ritonavir [19], chloroquine [10], or convalescent plasma [21].

Although the meta-analyzed results on the use of remdesivir may seem encouraging, its use in low-resource countries can be complicated by the cost of this drug. So, it can be assumed that up to now, the only drug with a large sample and that has demonstrated effectiveness has been dexamethasone, based on clinical trials conducted by the RECOVERY Collaborative Group and Tomazini [20,22]. Although only one of these two studies has reported adverse events, their conclusions are encouraging, mainly due to its low cost and easy accessibility in low-resource settings, as could occur in Latin American countries. This result is similar to that reported by Siemieniuk in a review published a few months ago [27]; this would indicate that studies may continue to produce relevant results for low-resource countries until a vaccine is available.

Among the limitations of this study, we can mention the rapid generation of new knowledge in times of the pandemic, which can potentially affect the timeliness of this review in a short time. Another limitation is the heterogeneity shown in the reviewed studies and their high risk of bias, which continues to affect the quality of the recommendations. In this review, we chose not to issue recommendations with the GRADE methodology, due to heterogeneity and high risk of bias.

Among the strengths of this study, focusing solely on inpatient studies, allowed us to review a larger volume of outcomes in these studies. The analysis of the main treatments proposed for hospitalized patients is of vital importance to reduce mortality in low-income countries; since the COVID-19 pandemic had an economic impact worldwide with the loss of jobs and economic decline [31] in countries with scarce resources. In these settings, the use of dexamethasone may be an affordable option for these countries. While there is no vaccine available, social distancing is so far the most crucial measure in controlling the spread of the disease [5].

In conclusion, dexamethasone would have a better result in hospitalized patients, although a detailed report of its adverse events is necessary. In Latin American countries, it is necessary to wait for the conclusion of some studies in the recruitment phase (3 clinical trials in Argentina and 1 in Mexico).

## Data Availability

The articles included in this systematic review are public.

## Authorship

RAAZ contributed to the development of the research project.

SCM, GFA and RAMGV performed data collection, analyzed, and interpreted the results.

RAAZ, SCM and RAMGV wrote the article. All authors reviewed and approved the final version.

Clinical effectiveness of drugs in hospitalized patients with COVID-19

Clinical effectiveness of drugs in hospitalized patients with COVID-19 infection: a systematic review and meta-analysis.

